# Multimorbidity and cardiometabolic clustering among university employees: findings from a comprehensive health assessment program in India

**DOI:** 10.64898/2026.10.20.26365860

**Authors:** Abhijeet Jagtap, Archana Ingole Patil, Abhay Kudale

## Abstract

**Objective:** To estimate the burden and patterns of multimorbidity and cardiometabolic multimorbidity among university employees undergoing a comprehensive health assessment programme, and to examine demographic and behavioural factors associated with multimorbidity.

**Design:** Cross-sectional study.

**Setting:** A public university in Maharashtra, India.

**Participants:** A total of 266 university employees who underwent a comprehensive health assessment were included.

**Main outcome measures:** Multimorbidity was defined as the presence of two or more of nine chronic conditions: hypertension, diabetes mellitus, dyslipidaemia, ischaemic heart disease, chronic kidney disease, thyroid disease, stroke, chronic obstructive pulmonary disease and cancer. Cardiometabolic multimorbidity was defined as the presence of two or more of hypertension, diabetes mellitus, dyslipidaemia, ischaemic heart disease and chronic kidney disease.

**Results:** Hypertension was identified in 119 participants (44.7%), comprising 50 participants with pre-existing hypertension and 69 newly identified during the assessment. Dyslipidaemia was present in 189 (71.1%) and diabetes mellitus in 45 (16.9%). Overall, 108 participants (40.6%) had multimorbidity and 103 (38.7%) had cardiometabolic multimorbidity. The prevalence of multimorbidity increased progressively with age, from 17.5% among participants aged <30 years to 83.3% among those aged ≥60 years. The predominant exact multimorbidity patterns were hypertension with dyslipidaemia alone (49 participants) and hypertension, diabetes mellitus and dyslipidaemia (27 participants). Overall, 173 participants (65.0%) had at least one newly identified hypertension, diabetes mellitus or dyslipidaemia diagnosis. In multivariable logistic regression, age (adjusted OR [aOR] 1.058 per year, 95% CI 1.027 to 1.091), BMI (aOR 1.163 per kg/m^2^, 95% CI 1.076 to 1.257) and tobacco use (aOR 3.296, 95% CI 1.377 to 7.888) were independently associated with multimorbidity. The model had an area under the receiver operating characteristic curve of 0.731.

**Conclusion:** Multimorbidity affected approximately two in five university employees, with cardiometabolic conditions accounting for most of the observed disease clustering. Comprehensive health assessment identified a substantial burden of previously unrecognised chronic disease. Integrating institutional or occupational health assessment with longitudinal primary care follow-up may provide an opportunity for earlier detection, risk-factor modification and coordinated management of multimorbidity.

## Introduction

Multimorbidity, generally defined as the coexistence of two or more chronic health conditions in the same individual, has become an increasingly important challenge for primary care. Its reported prevalence varies substantially according to the population studied, the conditions included and the methods used to ascertain disease, making consistent definitions and transparent measurement particularly important.^1^□^2^ Multimorbidity is clinically relevant because multiple chronic conditions can increase healthcare needs, complicate treatment decisions and require coordinated, person-centred care rather than disease-specific management delivered in isolation.^3^

The burden of multimorbidity is particularly important in India, where demographic transition, urbanisation and the increasing prevalence of non-communicable diseases have altered the health profile of working-age and older populations. Recent systematic and scoping reviews indicate substantial heterogeneity in the prevalence and definition of multimorbidity research in India, with cardiovascular, endocrine-metabolic and other chronic conditions frequently contributing to observed disease clusters. National data have also demonstrated a substantial burden of cardiometabolic risk factors and non-communicable diseases in the Indian population. These findings reinforce the importance of approaches that identify coexisting chronic conditions rather than considering each condition independently.

Primary care is particularly suited to the management of multimorbidity because it provides longitudinal, comprehensive and coordinated care across multiple health needs. Integrated approaches that combine risk assessment, early identification, treatment and follow-up are increasingly emphasized in primary healthcare frameworks. Evidence from primary care and community settings suggests that interventions for people with multimorbidity are most relevant when they address coordination of care, clinical priorities and the broader context in which patients manage chronic disease.^3^

Workplaces provide an additional opportunity for identifying chronic disease among adults who may otherwise have limited contact with preventive health services. Workplace-based health programs can facilitate cardiovascular risk assessment and identification of modifiable risk factors, although their effectiveness depends on linkage between detection and subsequent clinical management. In India, however, much of the available multimorbidity literature has focused on community-dwelling or older populations, with comparatively less attention to organized working populations and institutional health assessment programs.

Against this background, the present study examined the burden and patterns of multimorbidity among employees of a public university in India who participated in a comprehensive annual health assessment programme. The study specifically estimated the prevalence of multimorbidity and cardiometabolic multimorbidity, described predominant disease combinations and age-related patterns, quantified the contribution of newly identified chronic conditions, and examined demographic and behavioural factors associated with multimorbidity. The study was designed to provide a primary-care-oriented description of the chronic disease burden identified through an organised workplace health assessment programme and to examine its implications for continuity of care and community health practice.

## Methods

### Study design and setting

This was a cross-sectional analytical study based on data generated through an annual comprehensive health assessment programme conducted among employees of state university. The health assessment was conducted over two weeks in May 2025 as part of university community outreach activities supported through a Pradhan Mantri Uchchatar Shiksha Abhiyan (PM USHA) grant.

The study was reported in accordance with the Strengthening the Reporting of Observational Studies in Epidemiology (STROBE) recommendations.^9^ Statistical reporting and analysis were undertaken with reference to established SAMPL recommendations.^10^

### Study population and recruitment

All 306 university employees were invited to participate through an official institutional circular. Of these, 266 employees attended the health assessment, giving a participation proportion of 86.9%. The remaining 40 employees were unable to attend during the scheduled assessment period for routine administrative or personal reasons, including leave, deputation, summer vacation and other work-related commitments.

There was no a priori sample size calculation because the analytical sample comprised employees who participated in the scheduled institutional health assessment programme. The final analysis therefore included all 266 participants for whom the relevant health assessment and disease-status information were available.

### Ethical considerations

The study was approved by the Institutional Ethics Committee (Approval No. SPPU/IEC/2019/07). Written informed consent was obtained from participants before the health assessment and before use of their health information for research purposes.

### Health assessment and data collection

The assessment incorporated demographic characteristics, occupational information, lifestyle factors, anthropometric measurements, blood pressure measurements and biochemical investigations. Information on existing chronic diseases was obtained from the participant’s medical history and available health information.

### Blood pressure measurement and ascertainment of hypertension

Blood pressure was assessed as part of the health examination and hypertension was classified according to the Indian Society of Hypertension position statement.^11^ Blood pressure was measured by trained nursing personnel using a validated automated blood pressure monitor (OMRON HBP-1300). Participants were instructed to avoid smoking, caffeine consumption and strenuous physical activity before measurement. Following a five-minute seated rest period, three blood pressure readings were obtained at five-minute intervals, with participants seated comfortably and their arms supported at heart level. The average of the three readings was used for analysis.

Employees with no prior history of hypertension who had systolic blood pressure ≥140 mmHg and/or diastolic blood pressure ≥90 mmHg during the initial screening were invited to attend three subsequent follow-up visits on alternate days. Blood pressure was measured using the same standardised protocol at each visit. Employees without a prior history of hypertension were classified as having newly detected hypertension only when the blood pressure readings at all three follow-up visits remained at or above the threshold of 140 mmHg systolic and/or 90 mmHg diastolic.

Employees without a prior diagnosis whose blood pressure remained below this threshold were classified as normotensive. All employees identified with elevated blood pressure were counselled and referred for further medical evaluation and management.

### Diagnosis and classification of diabetes mellitus

Glycated haemoglobin (HbA1c) was measured using EDTA-anticoagulated whole blood by automated high-performance liquid chromatography (HPLC) using the D-10 analyser (Bio-Rad, USA). HbA1c was reported as a percentage. In accordance with the Indian Council of Medical Research (ICMR) Guidelines for Management of Type 2 Diabetes Mellitus (2018), an HbA1c value of ≥6.5% was considered indicative of diabetes mellitus.^12^ Diabetes mellitus was assessed using glycated haemoglobin (HbA1c), with a value of ≥6.5% (48 mmol/mol) considered indicative of diabetes, in accordance with the Indian Council of Medical Research (ICMR) Guidelines for Management of Type 2 Diabetes Mellitus, 2018. Participants with a previously documented diagnosis of diabetes were classified as having known diabetes. Those without a previous diagnosis who had an HbA1c value ≥6.5% on the single measurement performed during the health assessment were classified as having newly identified diabetes. As confirmatory testing was not performed, these cases were classified based on the screening measurement and were not considered independently confirmed clinical diagnoses. Previously known and newly identified cases were combined to estimate the overall prevalence of diabetes mellitus.

### Lipid profile assessment and classification of dyslipidaemia

The serum lipid profile was analysed using a fully automated biochemical analyser (COBAS C111 / MERCK LAB 300). The measured parameters included total cholesterol, triglycerides, high-density lipoprotein cholesterol (HDL-C), low-density lipoprotein cholesterol (LDL-C), very-low-density lipoprotein cholesterol (VLDL-C), total cholesterol-to-HDL-C ratio, and LDL-C-to-HDL-C ratio. The laboratory-reported reference limits were total cholesterol <190 mg/dL, triglycerides <150 mg/dL, HDL-C >50 mg/dL, LDL-C <100 mg/dL, VLDL-C <130 mg/dL, and total cholesterol-to-HDL-C ratio ≤5.00. Dyslipidaemia was classified as present when at least one measured lipid parameter fell outside its applicable laboratory reference range. These laboratory-based criteria were used consistently for the study analysis. These lab reference ranges were based on the cut offs recommended by the Lipid Association of India.^13^

Anthropometric assessment included body mass index (BMI), waist circumference, hip circumference and waist-to-hip ratio. Lifestyle variables included tobacco use, alcohol use and regular physical activity. Occupational variables included employment category and teaching/non-teaching status.

Other chronic conditions-ischemic heart disease (IHD), chronic kidney disease (CKD), stroke, cancer, chronic obstructive pulmonary disease (COPD) and thyroid disease—were ascertained solely from the history of present illness and past medical history recorded during the health assessment. No objective, biochemical, imaging or disease-specific investigations were undertaken specifically to screen for these conditions within the present study.

### Definition and ascertainment of chronic conditions

Participant-level disease variables were reconstructed by reconciling overlapping existing-disease and newly identified disease fields in the source dataset. Each participant was counted once for each chronic condition. Hypertension was defined as either pre-existing hypertension or newly identified hypertension during the assessment. Diabetes mellitus and dyslipidaemia were similarly classified by combining previously known disease with newly identified disease among participants without a corresponding previous diagnosis. The nine chronic conditions considered for the multimorbidity analysis were hypertension, diabetes mellitus, dyslipidaemia, IHD, CKD, thyroid disease, stroke, COPD and cancer.

Multimorbidity was defined as the presence of two or more of these nine chronic conditions. Because the predominant conditions in this dataset were cardiometabolic, a separate cardiometabolic multimorbidity variable was constructed as the presence of two or more of five conditions: hypertension, diabetes mellitus, dyslipidaemia, IHD and CKD. The disease ascertainment strategy was designed to avoid double counting where existing disease and newly identified disease were recorded in overlapping fields.

### Statistical analysis

Continuous variables were summarised using mean and SD when approximately normally distributed and median and IQR when appropriate. Categorical variables were summarised as frequencies and percentages. The prevalence of individual chronic conditions and multimorbidity was calculated for the complete analytical sample. The distribution of multimorbidity was examined across predefined age groups. Disease combinations were assessed using participant-level co-occurrence of chronic conditions. Pairwise co-occurrence counts were calculated as the number of participants having both conditions, with each participant contributing once to each relevant disease pair.

Univariable logistic regression was used to examine associations between selected demographic and behavioural factors and multimorbidity. A multivariable logistic regression model included age, BMI, sex, tobacco use, alcohol use and regular physical activity. Variables representing blood pressure, glucose, HbA1c or lipid measurements were not included as regression predictors because these measures contributed directly to the identification or definition of the cardiometabolic conditions constituting the multimorbidity outcome.

Associations are presented as ORs or adjusted ORs (aORs) with 95% CIs and p values. Model performance was additionally described using the area under the receiver operating characteristic curve (AUC), McFadden pseudo-R^2^ and the likelihood-ratio test. Statistical significance was considered at p<0.05.

All statistical analyses were performed using IBM SPSS Statistics, version 29.0 (IBM Corp., Armonk, NY, USA). Data preparation, participant-level disease-status reconciliation and derivation of composite variables were performed before the statistical analyses. Missingness was assessed for the variables included in the descriptive and regression analyses. All 266 participants had complete data for the multimorbidity outcome and the six predictors included in the final multivariable model.

## Results

### Participant characteristics

The analytical dataset comprised 266 university employees who attended the comprehensive health assessment programme. Of the 306 employees invited, 266 participated, giving a participation proportion of 86.9%. The remaining 40 employees were unable to attend during the scheduled assessment period because of routine administrative or personal circumstances, including leave, deputation, summer vacation and other work-related commitments.

The mean age of participants was 39.70±9.49 years, with a median age of 39 years (IQR 32–46). Men comprised 234 (88.0%) participants and women 32 (12.0%). Regular and contractual employees each contributed 133 (50.0%) participants, while 85 (32.0%) were teaching staff and 181 (68.0%) were non-teaching staff. Mean duration of service was 11.76±8.69 years.

The mean BMI was 25.09±3.81 kg/m^2^, with 138 (51.9%) participants having BMI ≥25 kg/m^2^. Tobacco use was reported by 35 (13.2%) participants and alcohol use by 37 (13.9%). Regular exercise was reported by 202 (75.9%), while 150 (56.4%) reported a family history of non-communicable disease. Detailed participant characteristics are presented in **Table 1**.

**Table 1.** Participant characteristics of the study population (N=266)

| **Characteristic** | **n (%) or mean± SD** |
| --- | --- |
| Age, years | 39.70±9.49 |
| Median age, years (IQR) | 39 (32–46) |
| **Age group, years** |  |
| <30 | 40 (15.0) |
| 30–39 | 99 (37.2) |
| 40–49 | 82 (30.8) |
| 50–59 | 39 (14.7) |
| ≥60 | 6 (2.3) |
| **Gender** |  |
| Male | 234 (88.0) |
| Female | 32 (12.0) |
| **Nature of service** |  |
| Regular | 133 (50.0) |
| Contractual | 133 (50.0) |
| **Cadre** |  |
| Teaching | 85 (32.0) |
| Non-teaching | 181 (68.0) |
| Duration of service, years | 11.76±8.69 |
| BMI, kg/m<sup>2</sup> | 25.09±3.81 |
| BMI ≥25 kg/m<sup>2</sup> | 138 (51.9) |
| Waist circumference, cm | 93.41±8.59 |
| Hip circumference, cm | 99.27±8.59 |
| Tobacco use | 35 (13.2) |
| Alcohol use | 37 (13.9) |
| Regular exercise | 202 (75.9) |
| Family history of NCD | 150 (56.4) |

(Values are mean± SD unless otherwise indicated. BMI, body mass index; IQR, interquartile range; NCD, non-communicable disease; SD, standard deviation.)

### Chronic-condition burden

Dyslipidaemia was the most frequently identified chronic condition, affecting 189 (71.1%) participants, followed by hypertension in 119 (44.7%) and diabetes mellitus in 45 (16.9%). Among the 119 participants classified as having hypertension, 50 had pre-existing hypertension and 69 were newly identified during the assessment. Similarly, 18 participants had pre-existing diabetes and 27 were newly identified, while 50 had pre-existing dyslipidaemia and 139 were newly identified.

Ischaemic heart disease was recorded in 8 (3.0%) participants, thyroid disease in 11 (4.1%), and both chronic kidney disease and stroke in 3 (1.1%) participants each. No participants were classified as having COPD or cancer.

The prevalence and ascertainment of individual chronic conditions are shown in **Table 2**.

**Table 2.**
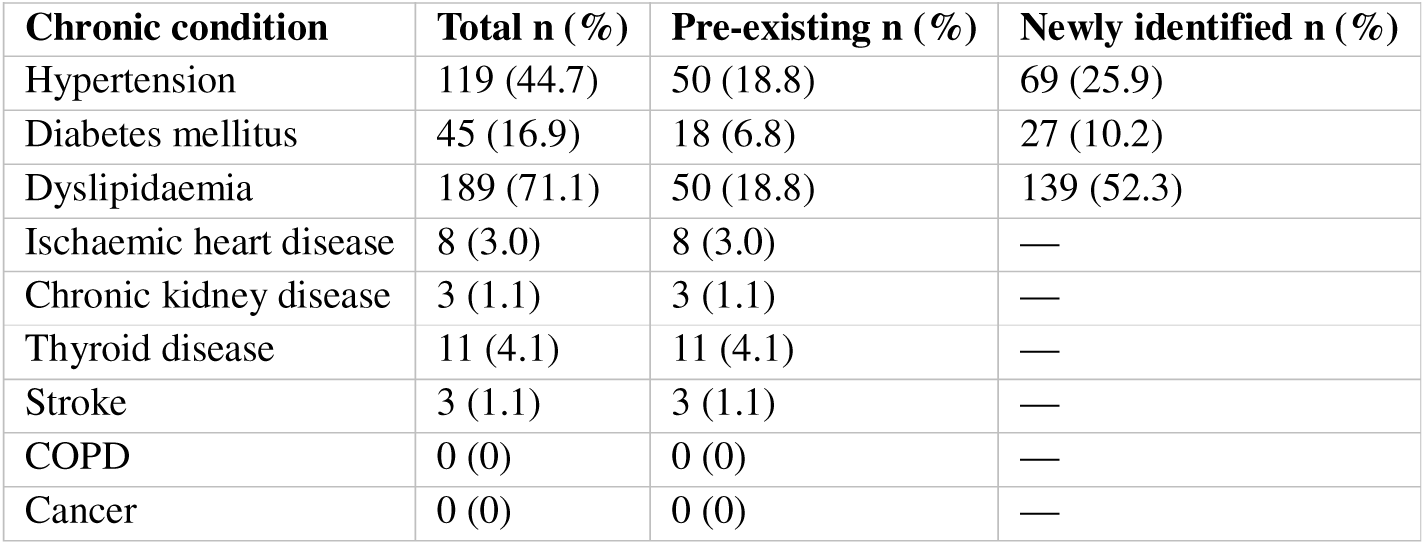
Prevalence and ascertainment of chronic conditions (N=266)

Newly identified disease was assessed only for hypertension, diabetes mellitus and dyslipidaemia. Other conditions were ascertained from medical history and therefore were not classified as newly identified through systematic disease-specific screening.

### Multimorbidity burden

Overall, 108 of 266 participants (40.6%) had multimorbidity, defined as the presence of at least two of the nine prespecified chronic conditions. The mean number of chronic conditions per participant was 1.42.

Forty-six participants (17.3%) had no recorded chronic condition, 112 (42.1%) had one condition, 65 (24.4%) had two, 37 (13.9%) had three, five (1.9%) had four and one (0.4%) had five conditions.

Thus, 103 participants (38.7%) had cardiometabolic multimorbidity, defined as the presence of at least two of hypertension, diabetes mellitus, dyslipidaemia, IHD or CKD.

The distribution of the number of chronic conditions, overall multimorbidity and cardiometabolic multimorbidity is presented in **Table 3**.

**Table 3.**
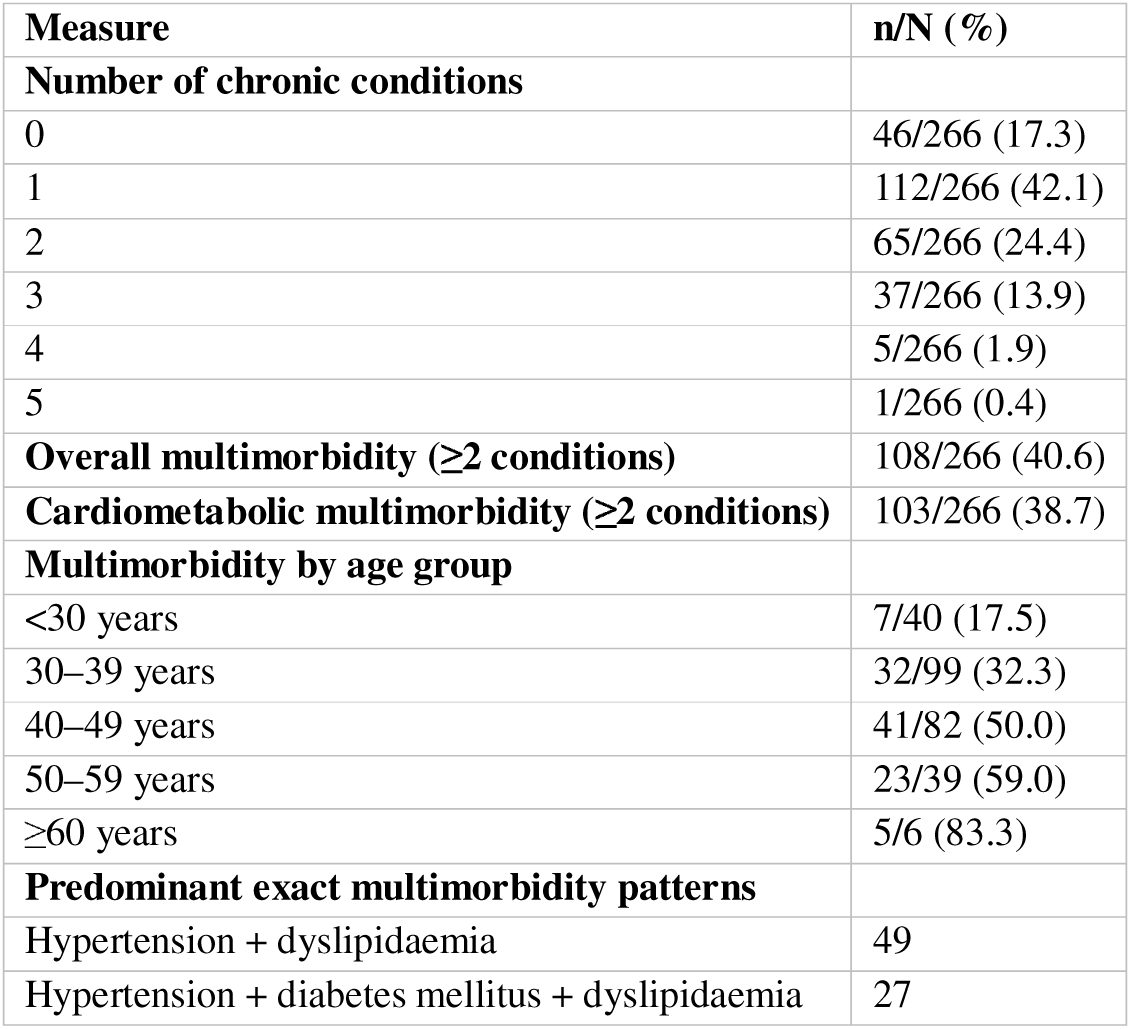
Multimorbidity burden, age gradient and predominant disease combinations.

| Measure | n/N (%) |
| --- | --- |
| **Number of chronic conditions** |  |
| 0 | 46/266 (17.3) |
| 1 | 112/266 (42.1) |
| 2 | 65/266 (24.4) |
| 3 | 37/266 (13.9) |
| 4 | 5/266 (1.9) |
| 5 | 1/266 (0.4) |
| **Overall multimorbidity (≥2 conditions)** | 108/266 (40.6) |
| **Cardiometabolic multimorbidity (≥2 conditions)** | 103/266 (38.7) |
| **Multimorbidity by age group** |  |
| <30 years | 7/40 (17.5) |
| 30–39 years | 32/99 (32.3) |
| 40–49 years | 41/82 (50.0) |
| 50–59 years | 23/39 (59.0) |
| ≥60 years | 5/6 (83.3) |
| **Predominant exact multimorbidity patterns** |  |
| Hypertension + dyslipidaemia | 49 |
| Hypertension + diabetes mellitus + dyslipidaemia | 27 |

The prevalence of multimorbidity increased progressively with age, from 17.5% among participants aged <30 years to 83.3% among those aged ≥60 years. (Ref. Fig.1)

**Figure 1.**
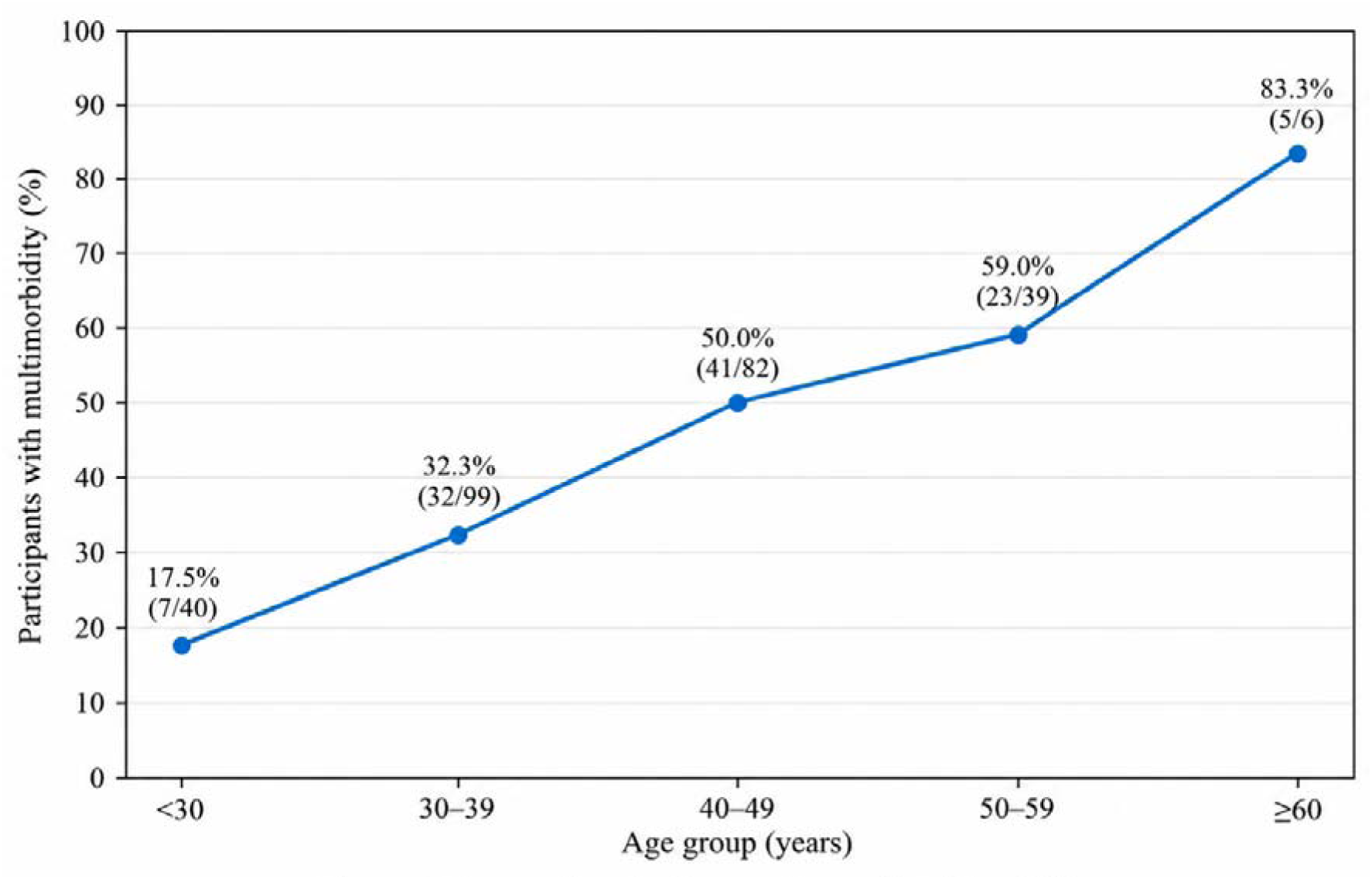
Age gradient in the prevalence of multimorbidity.

### Patterns of chronic-condition clustering

The most frequent exact multimorbidity pattern was the combination of hypertension and dyslipidaemia, observed in 49 participants. The combination of hypertension, diabetes mellitus and dyslipidaemia was observed in 27 participants. Other combinations involved thyroid disease, IHD, CKD and stroke in smaller numbers.

In pairwise co-occurrence analyses, which count participants with both conditions regardless of whether additional conditions were present, the most frequent pair was hypertension with dyslipidaemia (90 participants), followed by diabetes mellitus with dyslipidaemia (45) and hypertension with diabetes mellitus (32). Other observed pairwise combinations included dyslipidaemia with thyroid disease (8), dyslipidaemia with IHD (8), hypertension with thyroid disease (6), hypertension with IHD (6), diabetes mellitus with IHD (5), hypertension with CKD (3) and dyslipidaemia with stroke (3). The pairwise co-occurrence structure is displayed in **Figure 2**.

**Figure 2.**
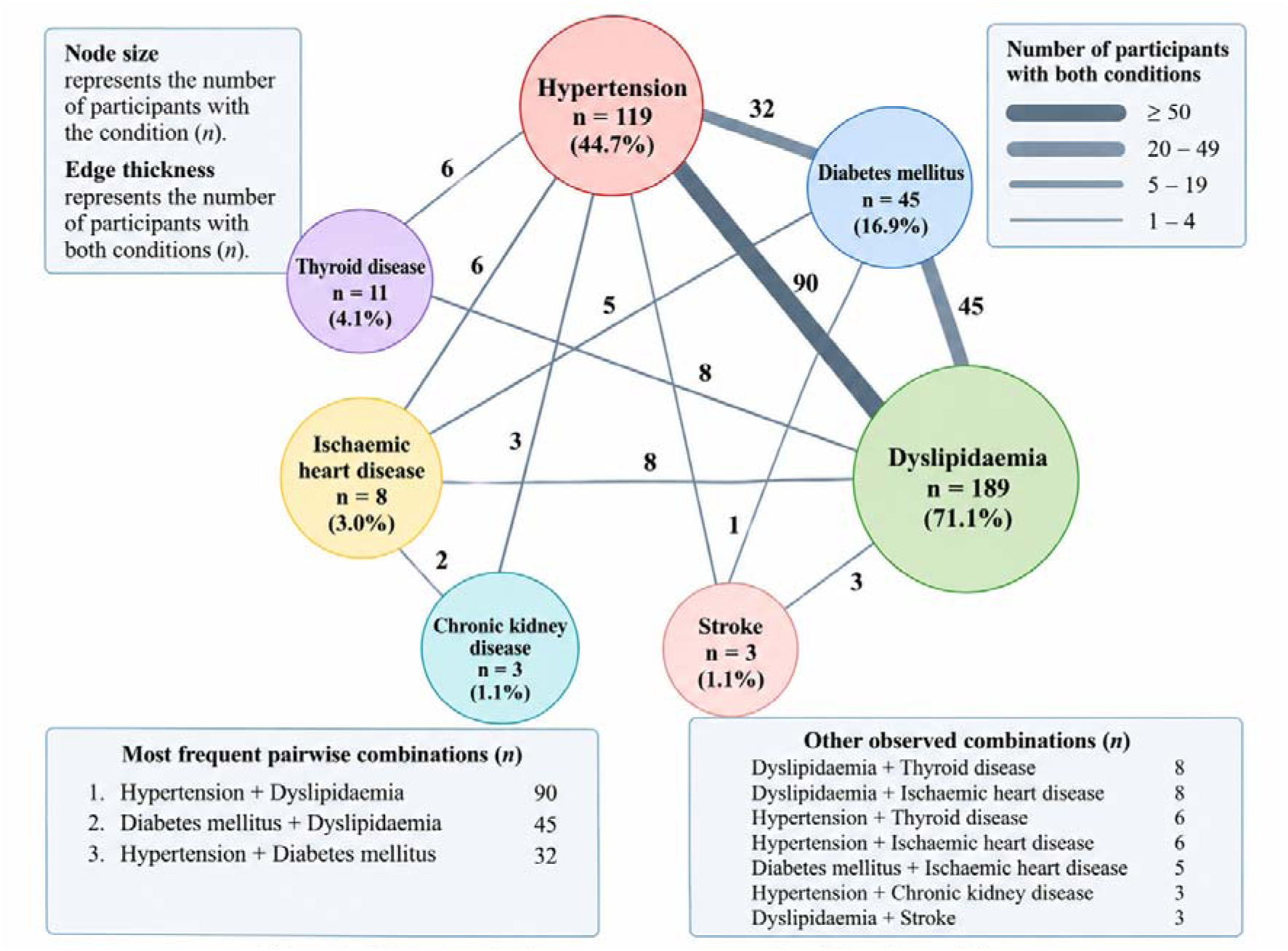
Observed pairwise co-occurrence network of chronic conditions. Nodes represent individual chronic conditions, with node size proportional lo the number of participants with each condition (*n,* %). Edges represent the number of participants with both conditions, with line thickness indicating the strength of co-occurence. Only observed co-occurrences are shown (*n* ≥ 1).

### Factors associated with multimorbidity

In univariable logistic regression, multimorbidity was associated with age ≥40 years, BMI ≥25 kg/m^2^ and tobacco use. Age ≥40 years was associated with higher odds of multimorbidity (OR 3.05, 95% CI 1.83 to 5.07; p<0.001), as was BMI ≥25 kg/m^2^ (OR 2.44, 95% CI 1.47 to 4.04; p<0.001) and tobacco use (OR 3.29, 95% CI 1.56 to 6.95; p=0.002). Alcohol use was not statistically associated with multimorbidity (OR 1.89, 95% CI 0.94 to 3.79; p=0.076). No evidence of an association was observed for male sex or lack of regular exercise.

In the multivariable model including age, BMI, sex, tobacco use, alcohol use and regular exercise, age, BMI and tobacco use remained associated with multimorbidity. Each additional year of age was associated with higher adjusted odds of multimorbidity (aOR 1.058, 95% CI 1.027 to 1.091; p<0.001). Each 1 kg/m^2^ increase in BMI was associated with higher adjusted odds (aOR 1.163, 95% CI 1.076 to 1.257; p<0.001). Tobacco use was also associated with higher adjusted odds of multimorbidity (aOR 3.296, 95% CI 1.377 to 7.888; p=0.007). Male sex, alcohol use and lack of regular exercise were not statistically associated with multimorbidity after adjustment.

The multivariable model had an area under the receiver operating characteristic curve of 0.731, a McFadden pseudo-R^2^ of 0.125 and a likelihood-ratio test p<0.001. The complete univariable and multivariable estimates are presented in **Table 4**, with adjusted associations shown graphically in **Figure 3**.

**Figure 3.**
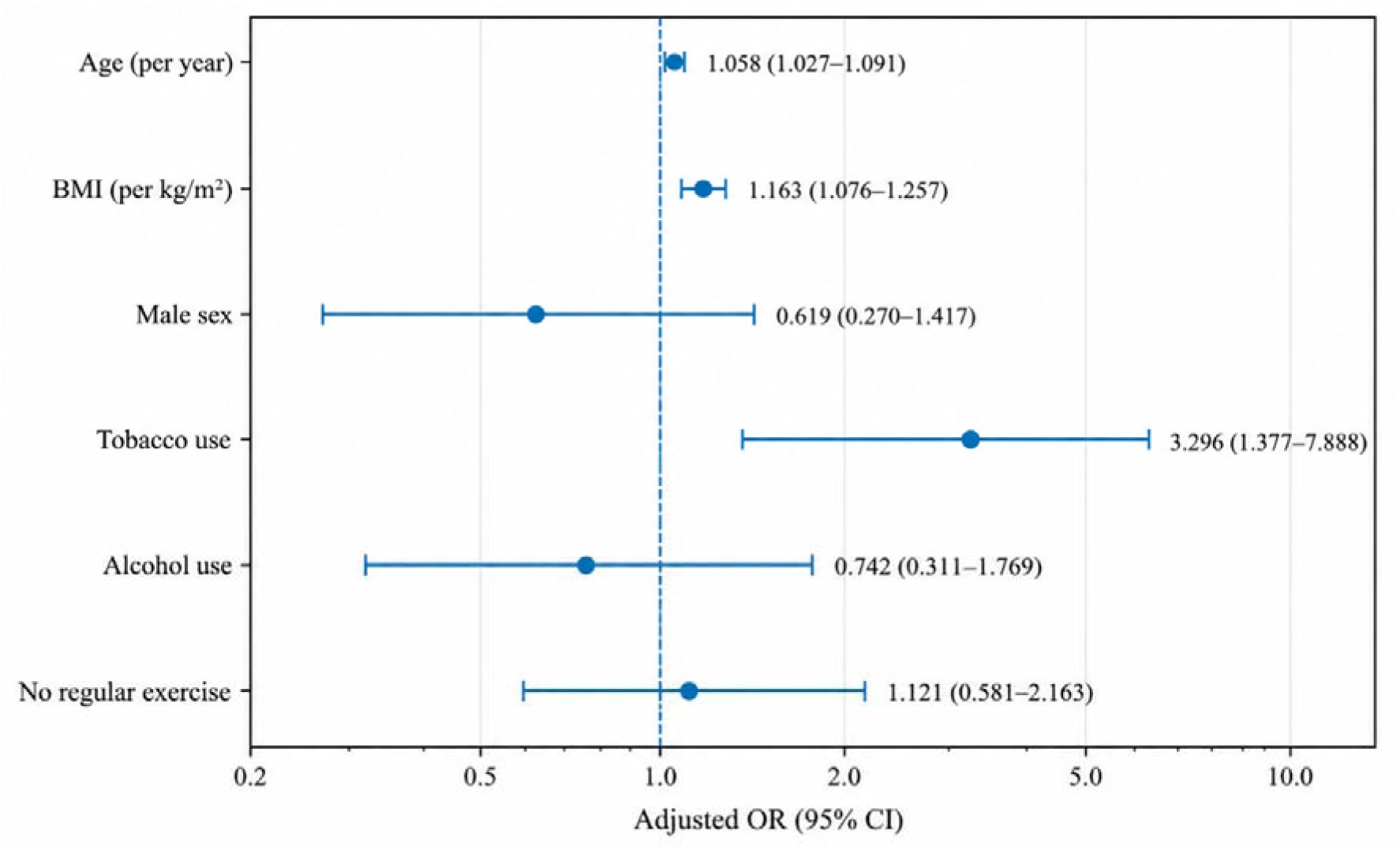
Forest plot of multivariable logistic regression associations with multimorbidity.

**Table 4.** Univariable and multivariable logistic regression analysis of factors associated with multimorbidity.

| Predictor | Crude OR | 95% CI | p value | Adjusted OR | 95% CI | p value |
| --- | --- | --- | --- | --- | --- | --- |
| Age, per year | — | — | — | 1.058 | 1.027–1.091 | <0.001 |
| Age ≥40 years | 3.05 | 1.83–5.07 | <0.001 | — | — | — |
| BMI, per kg/m<sup>2</sup> | — | — | — | 1.163 | 1.076–1.257 | <0.001 |
| BMI ≥25 kg/m<sup>2</sup> | 2.44 | 1.47–4.04 | <0.001 | — | — | — |
| Male sex | 0.75 | 0.36–1.57 | 0.442 | 0.619 | 0.270–1.417 | 0.256 |
| Tobacco use | 3.29 | 1.56–6.95 | 0.002 | 3.296 | 1.377–7.888 | 0.007 |
| Alcohol use | 1.89 | 0.94–3.79 | 0.076 | 0.742 | 0.311–1.769 | 0.501 |
| No regular exercise | 0.84 | 0.47–1.50 | 0.562 | 1.121 | 0.581–2.163 | 0.733 |

(OR, odds ratio; aOR, adjusted odds ratio; CI, confidence interval; BMI, body mass index.)

### Newly identified cardiometabolic chronic disease

A total of 173 participants (65.0%) had at least one newly identified hypertension, diabetes mellitus or dyslipidaemia diagnosis during the comprehensive health assessment.

Among the 119 participants classified as having hypertension, 69 (58.0%) were newly identified. Of the 45 participants with diabetes mellitus, 27 (60.0%) were newly identified. Among the 189 participants with dyslipidaemia, 139 (73.5%) were newly identified.

Thus, newly identified disease accounted for a substantial proportion of the total burden of the three objectively assessed cardiometabolic conditions. The high proportion of previously unrecognized cardiometabolic disease identified during the assessment is an important finding (**figure 4**).

**Figure 4.**
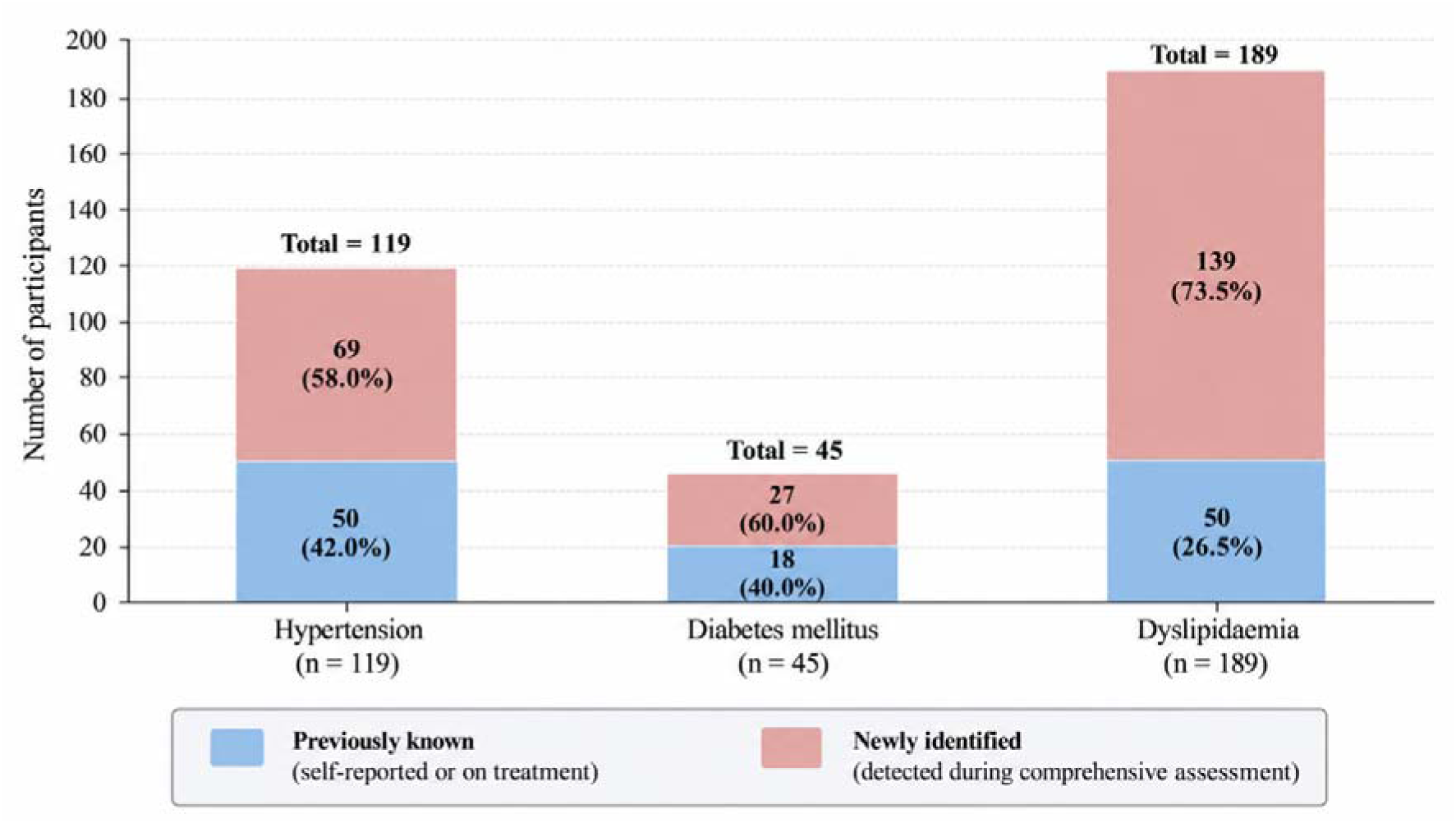
Previously known versus newly identified cardiometabolic disease among study participants (N = 266) **Note:** Percentages are calculated within each disease category. A substantial proportion of hypertension (58.0%), diabetes mellitus (60.0%) and dyslipidaemia (73.5%) were newly identified during the workplace health assessment programme.

## Discussion

### Principal findings

This study identified a substantial burden of multimorbidity among university employees participating in a comprehensive institutional health assessment programme. Overall, 108 of 266 participants (40.6%) had two or more of the nine chronic conditions considered, while 103 (38.7%) met the study definition of cardiometabolic multimorbidity. The prevalence of multimorbidity increased markedly with age, from 17.5% among participants aged below 30 years to 83.3% among those aged 60 years or older. Hypertension, dyslipidaemia and diabetes mellitus were the predominant conditions, and combinations involving these cardiometabolic conditions accounted for much of the observed disease clustering.

An important finding was the substantial burden of previously unrecognised disease. At least one newly identified finding related to hypertension, diabetes mellitus or dyslipidaemia was recorded in 173 participants (65.0%). Among participants classified as having these respective conditions, 58.0% of hypertension cases, 60.0% of diabetes cases and 73.5% of dyslipidaemia cases were newly identified during the assessment. These findings highlight the potential of organised workplace health assessments to identify unmet diagnostic needs among employed adults. However, newly identified cases should not be interpreted as incident disease, as the cross-sectional assessment cannot establish when the underlying abnormalities first developed.

In the multivariable analysis, age, body mass index (BMI) and tobacco use were independently associated with multimorbidity. The adjusted odds increased by approximately 6% for each additional year of age (adjusted OR [aOR] 1.058, 95% CI 1.027 to 1.091). Higher BMI was also associated with multimorbidity (aOR 1.163 per kg/m^2^, 95% CI 1.076 to 1.257), as was tobacco use (aOR 3.296, 95% CI 1.377 to 7.888). The model had an area under the receiver operating characteristic curve of 0.731, indicating moderate discriminatory performance in this study population. Taken together, the findings suggest that multimorbidity is already relevant in a working-age population and that integrated assessment may identify coexisting conditions before they are all clinically recognised.

### Cardiometabolic clustering and multimorbidity

The predominance of cardiometabolic conditions was a central finding. Dyslipidaemia was present in 189 participants (71.1%), hypertension in 119 (44.7%) and diabetes mellitus in 45 (16.9%). The most frequent pairwise combinations were hypertension with dyslipidaemia (n=90), diabetes mellitus with dyslipidaemia (n=45) and hypertension with diabetes mellitus (n=32). When exact disease patterns were examined, hypertension with dyslipidaemia alone was recorded in 49 participants, while 27 had the combination of hypertension, diabetes mellitus and dyslipidaemia. These patterns demonstrate that chronic conditions in this population often occurred together rather than as isolated diagnoses.

The findings are consistent with Indian research showing a substantial burden of cardiometabolic disease and the frequent coexistence of risk factors. The ICMR–INDIAB national study of metabolic non-communicable diseases documented the considerable burden of metabolic abnormalities across India. More recently, the ICMR–INDIAB-25 study, based on lipid profiles from 23,665 adults, reported a weighted dyslipidaemia prevalence of 87.3%, with low high-density lipoprotein cholesterol (HDL-C) being the most common lipid abnormality, affecting 66.8% of participants. Dyslipidaemia was more prevalent among people with adverse glycaemic status, higher BMI and hypertension, although it was also observed in a substantial proportion of individuals without diabetes, hypertension or obesity.^23^ These national findings reinforce the importance of considering lipid abnormalities alongside blood pressure, glycaemic status and adiposity when assessing cardiometabolic risk.

The dyslipidaemia prevalence of 71.1% in the present study should not, however, be directly equated with the 87.3% reported by ICMR–INDIAB-25. The studies differed in sampling design, population composition and operational definitions. ICMR–INDIAB-25 used specified Lipid Association of India thresholds for total cholesterol, triglycerides, LDL-C and HDL-C, whereas the present study classified dyslipidaemia using the applicable laboratory reference ranges used during the institutional health assessment.^23^ The estimates are therefore not directly interchangeable. Nevertheless, both studies point to a considerable burden of lipid abnormalities and support the value of including lipid assessment in comprehensive cardiometabolic evaluation.

The high proportion of dyslipidaemia in the present study also needs to be interpreted in light of how the conditions were identified. Of the 189 participants classified as having dyslipidaemia, 139 (73.5%) had newly identified abnormalities, compared with 50 with pre-existing dyslipidaemia. The prevalence therefore reflects both the underlying burden of lipid abnormalities and the ability of biochemical assessment to identify findings that had not previously been recognised. Laboratory-defined abnormalities identified during a screening assessment are important for case finding, but their clinical interpretation and management should take account of the specific lipid components, overall cardiovascular risk and appropriate clinical confirmation or follow-up.

The observed multimorbidity prevalence is also broadly consistent in magnitude with evidence from Indian population studies, although direct comparisons require caution. Pattanaik and colleagues analysed data from 7,118 adults aged 50 years or older in the Study on Global Ageing and Adult Health (SAGE) Wave 2 and reported a multimorbidity prevalence of 45.14%. Their analysis considered eight self-reported chronic conditions, including hypertension, diabetes, angina, stroke, arthritis, asthma, chronic lung disease and depression. Hypertension featured prominently in the observed disease combinations.^2^□ The prevalence reported in that study is numerically similar to the 40.6% observed in our university workforce, but the two estimates represent different populations and measurement approaches. The SAGE analysis was restricted to older adults and relied on self-reported conditions, whereas the present study included a predominantly working-age population and incorporated clinical and biochemical assessment of key cardiometabolic conditions. The difference in the conditions included in the multimorbidity definition also limits direct comparison.

Taken together, these findings support an integrated approach to chronic disease assessment. Evaluating hypertension, diabetes and lipid abnormalities independently may overlook the extent to which these conditions coexist in the same individuals. A combined assessment of blood pressure, glycaemic status, lipid profile, BMI and behavioural risk factors can provide a more complete picture of cardiometabolic burden and help identify individuals who may benefit from comprehensive risk assessment and coordinated follow-up.

### Age, BMI and tobacco use

Age was independently associated with multimorbidity, and the adjusted odds increased by approximately 6% per year. The marked age gradient in the descriptive findings—from 17.5% among participants aged below 30 years to 83.3% among those aged 60 years or older—also indicates an accumulation of chronic conditions across adulthood. This pattern is consistent with previous Indian studies documenting higher multimorbidity prevalence among older adults and variation across age groups and disease combinations.^1^□–^1^□ The SAGE Wave 2 analysis similarly documented a substantial burden among adults aged 50 years or older.^2^□ The present study extends this evidence to an organised working population, demonstrating that multimorbidity is not confined to older adults or people outside the workforce.

BMI was also independently associated with multimorbidity. Each 1 kg/m^2^ increase in BMI was associated with higher adjusted odds of multimorbidity (aOR 1.163, 95% CI 1.076 to 1.257). Previous evidence, including a systematic review and dose-response meta-analysis, has identified higher BMI as an important correlate of multimorbidity.^1^□ Excess adiposity may contribute to the development or clustering of several chronic conditions, particularly cardiometabolic disorders. Nevertheless, the cross-sectional design of the present study does not establish temporal sequence or causation. The association supports the inclusion of anthropometric assessment and weight-related counselling within integrated primary care and workplace health programmes.

Tobacco use was also independently associated with multimorbidity (aOR 3.296, 95% CI 1.377 to 7.888). Tobacco exposure is relevant to multiple chronic disease pathways, making tobacco assessment and cessation support important components of comprehensive chronic disease prevention and management. The confidence interval around the estimate was relatively wide, so the magnitude of association should be interpreted cautiously. Sex, alcohol use and regular physical activity were not statistically associated with multimorbidity after adjustment. These findings should not be interpreted as evidence that these factors are clinically unimportant; the estimates may reflect the size and characteristics of this study population, including its predominantly male composition and the distribution of behavioural exposures.

### Previously unrecognised chronic disease and implications for primary care

The high proportion of newly identified cardiometabolic disease is one of the most important practical findings. Nearly two-thirds of participants had at least one newly identified finding related to hypertension, diabetes mellitus or dyslipidaemia. This distinction between the presence of a condition and its prior recognition within the healthcare system is important because people may remain unaware of abnormalities until they undergo systematic assessment.

The findings are particularly relevant to primary care, where detection should be linked to diagnostic confirmation when indicated, cardiovascular risk assessment, counselling, treatment or referral, and longitudinal follow-up. A screening result alone does not ensure improved health outcomes. The value of an assessment programme depends on whether participants with abnormal findings can access appropriate clinical evaluation and sustained management. Integrated approaches to multimorbidity are therefore particularly relevant because care must address multiple conditions and competing priorities rather than manage each diagnosis in isolation.^3^,□,^1^□

Workplaces and educational institutions may provide a useful setting for identifying chronic disease among adults who may not routinely access preventive services. Evidence on workplace health interventions suggests that organised programmes can address cardiovascular and other non-communicable disease risk factors, although their impact depends on implementation and the continuity of care after screening. □,^2^ □The present study suggests that a comprehensive institutional assessment can identify a considerable burden of previously unrecognised cardiometabolic abnormalities. Such programmes should complement routine primary care rather than function as a substitute for it.

### Implications for occupational health practice and workplace health programmes

The findings highlight the potential role of workplace health programmes in identifying and addressing the burden of multimorbidity among employed adults. In this study, 65.0% of participants had at least one newly identified finding related to hypertension, diabetes mellitus or dyslipidaemia, demonstrating that important cardiometabolic abnormalities may remain unrecognised even among individuals engaged in routine employment. Periodic workplace health assessments could therefore provide an opportunity to identify coexisting risk factors and chronic conditions through an integrated approach rather than through isolated, disease-specific screening.

Occupational health programmes should incorporate assessment of blood pressure, glycaemic status, lipid profile, BMI and relevant behavioural risk factors, alongside appropriate evaluation of existing chronic conditions. The observed associations of multimorbidity with age, higher BMI and tobacco use further support age-appropriate risk assessment, weight management and tobacco cessation as components of workplace health promotion. However, identification of an abnormality should be considered the beginning of care rather than its endpoint. Programmes should establish clear pathways for clinical evaluation, confirmatory testing where indicated, referral, treatment and follow-up. Such pathways are particularly important when screening identifies multiple abnormalities requiring coordinated management.

The findings also suggest that workplace health initiatives should extend beyond the immediate assessment period. Health promotion activities, periodic reassessment and mechanisms for monitoring whether employees have accessed recommended clinical care could help connect early identification with ongoing disease management. Although the present study did not evaluate the effectiveness of these interventions, it identifies a substantial burden of previously unrecognised cardiometabolic disease that warrants attention within institutional and occupational health planning. Workplace programmes should complement existing health services and facilitate access to appropriate clinical care rather than function as stand-alone screening activities.□,^2^□

### Integrating occupational health with primary care: implications for the prevention and management of multimorbidity

This study contributes to the understanding of multimorbidity by examining an organised working population in which cardiometabolic conditions accounted for much of the observed disease clustering. While multimorbidity research in India has frequently focused on older adults and community-based populations, the present findings demonstrate that substantial coexisting chronic disease can also be identified among employees, including those in the working-age population. The high prevalence of newly identified cardiometabolic abnormalities further highlights the distinction between the presence of disease and its prior recognition within the healthcare system. These findings complement national evidence on dyslipidaemia and population-based research on multimorbidity, while differences in study populations, condition definitions and ascertainment methods must be considered when interpreting comparisons.^23^,^2^□

A practical implication is the need for stronger integration between occupational health services and primary care. Workplace assessments can provide an initial opportunity to identify risk factors and possible chronic disease, whereas family medicine and primary care services are essential for clinical confirmation, individualised risk assessment, treatment decisions and continuity of care. Establishing referral and feedback mechanisms between these settings may help ensure that employees with abnormal findings receive appropriate evaluation and that those with multiple chronic conditions are not managed through disconnected, disease-specific pathways. Such coordination is consistent with the broader principles of person-centred, integrated care for multimorbidity.^3^,^1^□,^21^,^22^

The university setting may offer a useful platform for linking health assessment with sustained prevention and care. However, the present study was not designed to assess referral completion, treatment uptake, risk-factor control or long-term outcomes. Future prospective studies should evaluate whether integrated workplace–primary care pathways improve these outcomes, identify implementation barriers and determine whether the approach can be extended to other occupational settings. By connecting occupational health assessment with longitudinal primary care, institutions may be able to move beyond one-time detection towards a more coordinated approach to the prevention and management of multimorbidity among working adults.

### Strengths and limitations

This study has several strengths. The health assessment was offered to the entire university employee population rather than to a selected subgroup, and participation was high at 86.9%. The dataset included demographic, occupational, anthropometric, behavioural and biochemical information, enabling a multidimensional assessment of chronic disease burden. Participant-level reconciliation of existing and newly identified disease fields reduced the risk of double counting. In addition, the multivariable model did not include blood pressure, glucose, HbA1c or lipid measurements as independent predictors when these measures contributed to the identification of conditions constituting the multimorbidity outcome, reducing the risk of direct predictor–outcome overlap.

Several limitations should also be considered. First, the cross-sectional design prevents conclusions about temporality or causation. Second, the study was conducted at a single public university and should not be considered representative of all university employees or the wider Indian working population. Third, the population was predominantly male, limiting the precision and generalisability of sex-specific observations. Fourth, 40 invited employees did not attend the assessment, creating the possibility of non-participation bias if their health profiles differed from those of participants.

Fifth, ascertainment was not uniform across all chronic conditions. Hypertension, diabetes mellitus and dyslipidaemia were assessed through clinical or biochemical procedures, whereas several other chronic conditions were identified from present and past medical history without disease-specific investigations. This may have led to under-ascertainment of undiagnosed conditions such as chronic kidney disease, chronic obstructive pulmonary disease or thyroid disease. The greater use of objective assessment for cardiometabolic conditions may also have increased their detection relative to conditions ascertained from history alone.

Sixth, the dyslipidaemia prevalence depends on the laboratory reference ranges used for classification and is not directly comparable with estimates from studies applying other diagnostic thresholds. The ICMR–INDIAB-25 study, for example, used specified Lipid Association of India criteria and a nationally representative sampling design, whereas the present study used the reference ranges applied during the institutional health assessment.^23^ Finally, the nine-condition framework represents an operational definition based on the available data and does not include every possible chronic condition. These considerations should be taken into account when comparing the reported multimorbidity prevalence with estimates from studies using different populations, condition lists and ascertainment methods.

## Conclusion

Multimorbidity represents an important health burden among university employees, with cardiometabolic conditions forming the predominant pattern of coexisting chronic disease. The strong age gradient and independent associations with higher BMI and tobacco use highlight the importance of integrated risk assessment alongside routine chronic disease care.

A particularly important finding is the substantial burden of previously unrecognised cardiometabolic disease. Comprehensive workplace health assessment can therefore serve as an entry point for earlier identification, provided that abnormal findings are linked to appropriate clinical confirmation, risk-factor modification, treatment and longitudinal follow-up. The findings support a shift from isolated disease-specific screening towards integrated, multimorbidity-oriented assessment and continuity of care. Strengthening the connection between institutional health programmes and family medicine or primary care may provide a practical approach to the earlier identification and coordinated management of chronic disease in working populations.

## Data Availability

All data produced in the present study are available upon reasonable request to the authors

## Notes

### Competing Interest Statement

The authors have declared no competing interest.

### Author Declarations

The study was approved by the Institutional Ethics Committee of Savitribai Phule Pune University (Approval No. SPPU/IEC/2019/07).

